# Adverse Drug Events Across Data-Production Contexts: Multilingual Detection, Alignment, and Cross-Genre Discourse Analysis

**DOI:** 10.64898/2026.08.08.26360012

**Authors:** Yujun Ma, Davy Weissenbacher, Jake Patock, Graciela Gonzalez Hernandez

## Abstract

Adverse drug event (ADE) evidence is produced across patient-generated, clinical, and scientific settings that differ in language, documentation purpose, terminology, and degree of standardization. These differences shape both which adverse experiences become visible to pharmacovigilance systems and how readily they can be linked to curated drug-safety knowledge. We examine these relationships across five corpora representing distinct data-production settings: ADE Corpus V2 (medical case reports), SMM4H-2026 Task 1 (multi-lingual user-generated health content), CADEC V2 (patient-forum narratives), the Dutch ADE Corpus (EHR clinical notes), and TwiMed-PubMed (biomedical literature).

A shared BERTopic analysis of ADE-positive texts concerning antidepressants and antihypertensives across the four English-language corpora identified nine interpretable topics. CADEC V2 contained a more differentiated distribution of symptom-specific themes, including sexual effects, suicidal or panic-related thoughts, vivid dreams, and memory difficulties, whereas SMM4H-2026, TwiMed-PubMed, and ADE Corpus V2 were dominated by a broader medication, sleep, tiredness, and pain theme. These patterns indicate that data-production context shapes what adverse experiences are expressed and standardized, with patient-generated narratives surfacing subjective, symptom-specific experience largely absent from clinical and scientific sources.

We further show that this context shapes how readily real-world drug mentions can be linked to curated pharmacovigilance knowledge. Using SIDER 4.1 as a retrieval resource, we find substantial cross-corpus mismatches between real-world drug mentions and SIDER’s predominantly English, generic-name vocabulary: CADEC V2 achieved only 9.5% exact-match coverage, with unmatched mentions frequently involving brand names, misspellings, and language-specific variants, compared to 91.0% coverage in TwiMed-PubMed’s formally standardized biomedical literature.

To probe how these representational differences interact with automated detection, we compare corpus-specific QLoRA fine-tuning of Llama-3.2-3B with retrieval-augmented inference using Llama-3.1-70B and Llama-3.1-405B grounded in SIDER-retrieved evidence. QLoRA-Llama-3B achieved the highest micro-averaged F1 scores on ADE Corpus V2 (0.91), CADEC V2 (0.88), and SMM4H-2026 (0.80), whereas SIDER-grounded inference with Llama-3.1-405B achieved the highest scores on Dutch ADE (0.95) and TwiMed-PubMed (0.91); these corpus-dependent patterns should not be interpreted as a controlled comparison of adaptation strategies, since model scale, task formulation, and available supervision differ across datasets. Together, our findings indicate that data-production context influences what adverse experiences are expressed, how they are standardized, and how readily they can be retrieved and computationally detected. Pharmacovigilance systems should therefore combine source-sensitive supervision with external knowledge grounding while explicitly monitoring gaps between real-world language and curated drug-safety resources.

## 1. Introduction

Adverse drug events (ADEs) are harmful or unintended outcomes associated with medication use, including adverse reactions, medication errors, and other drug-related injuries.^1,2^ They remain a major patient-safety concern and are associated with increased mortality, prolonged hospitalization, elevated healthcare costs, disability, and death.^3,4^ Although clinical trials and spontaneous reporting systems provide important safety evidence, many ADEs are underreported or documented only indirectly in electronic health records (EHRs), patient-generated posts, medical forums, case reports, and biomedical literature.^5,6^ Automated methods can support earlier identification of such signals, though their effectiveness depends on how well they account for the representational differences across these sources.^7–9^

These sources are not interchangeable representations of the same pharmacovigilance evidence. They are produced for different audiences and purposes and vary in terminology, language, documentation practices, and degree of standardization. Clinical and scientific sources may explicitly describe drug–outcome relations using specialized terminology, whereas social-media posts and patient-forum narratives often contain misspellings, slang, multilingual expressions, implicit symptom descriptions, and extended accounts of lived experience.^10^ In this study, we use the term *data-production context* to capture these differences. Through cross-source topic modeling, we examine how such contexts shape which adverse experiences are expressed and how they are framed, and how readily the resulting drug mentions can be linked to standardized pharmacovigilance knowledge. We complement this analysis with ADE detection experiments, using them as a lens to further probe how source-specific supervision, pretrained model knowledge, and external evidence interact with these representational differences, rather than as an end in themselves.

Prior discourse-oriented work has largely examined patient narratives within a single platform or population,^11,12^ with relatively little attention to how ADE expression and framing compare across patient-generated, clinical, and scientific sources within a shared analytic frame. A related open question is the form of knowledge on which downstream detection systems rely.^13^ Fine-tuning learns from corpus-specific labeled examples, whereas retrieval-augmented generation (RAG) introduces external evidence at inference time. In this study, retrieval is grounded in SIDER, a curated resource containing standardized drug–adverse-effect associations derived primarily from drug labels and clinical-trial information.^14^ Although such knowledge can support ADE detection, SIDER’s predominantly English, standardized vocabulary may align unevenly with brand names, misspellings, colloquial forms, and language-specific drug mentions found in real-world text. Prior work has shown that SIDER-grounded RAG can improve drug–side-effect question answering on a single benchmark;^15^ however, its coverage across heterogeneous data-production contexts remains unclear. Recent LLM-based ADE detection work using zero- and few-shot prompting, parameter-efficient fine-tuning, and structured knowledge construction^16–18^ has likewise been evaluated largely on a single corpus or a small number of closely related sources,^19,20^ with limited attention to how data-production context shapes both ADE expression and model performance.

Taken together, these gaps motivate three research questions: (1) how ADE discourse differs across patient-generated, clinical, and scientific sources; (2) how well drug mentions from these settings can be linked to SIDER; and (3) how corpus-specific fine-tuning, prompting, and SIDER-grounded inference perform across different data-production contexts, as a means of probing the downstream consequences of (1) and (2).

Figure 1 summarizes the study design. As shown in Panel 1, we analyze five corpora representing distinct data-production contexts: ADE Corpus V2 medical case reports,^21^ SMM4H-2026 Task 1 multilingual user-generated health content,^22^ CADEC V2 patient-forum narratives,^23^ the Dutch ADE Corpus EHR clinical notes,^24^ and TwiMed-PubMed biomedical literature.^25^ Panel 2 presents the shared BERTopic analysis^26^ of ADE-positive English-language texts concerning antidepressants and antihypertensives from four corpora, which we use to examine how adverse experiences differ across data-production contexts and how they relate to SIDER coverage. Panel 3 presents the ADE detection approaches, including corpus-specific QLoRA fine-tuning of Llama-3.2-3B^27^ and SIDER-grounded inference with Llama-3.1-70B and Llama-3.1-405B, alongside zero- and few-shot prompting baselines, which together let us assess how readily real-world drug mentions from each corpus can be linked to the standardized external knowledge resource and how this alignment relates to detection outcomes.

**Fig. 1:**
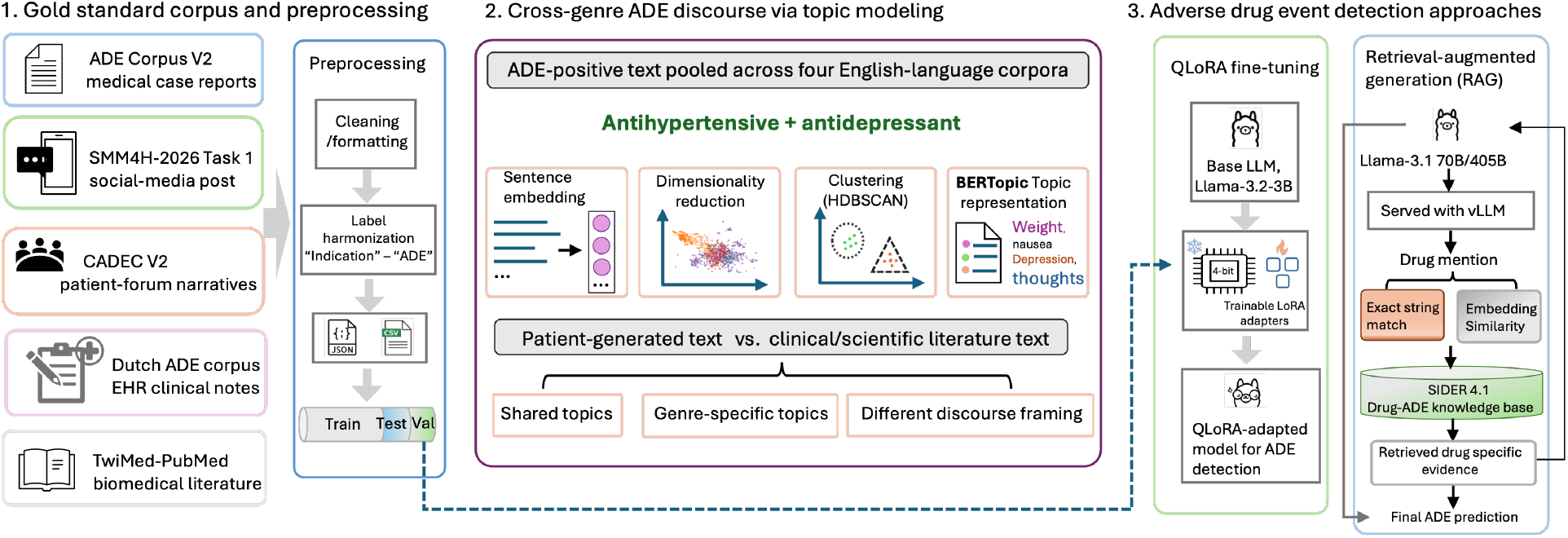
Overview of the study design. Five ADE corpora representing distinct healthcare data-production settings are standardized into a common binary ADE detection task. We first characterize cross-source differences in adverse-event discourse through BERTopic analysis of English-language corpora. We then compare source-specific fine-tuning with SIDER-grounded retrieval-augmented inference to examine how external pharmacological knowledge interacts with different documentation settings.

**Fig. 2:**
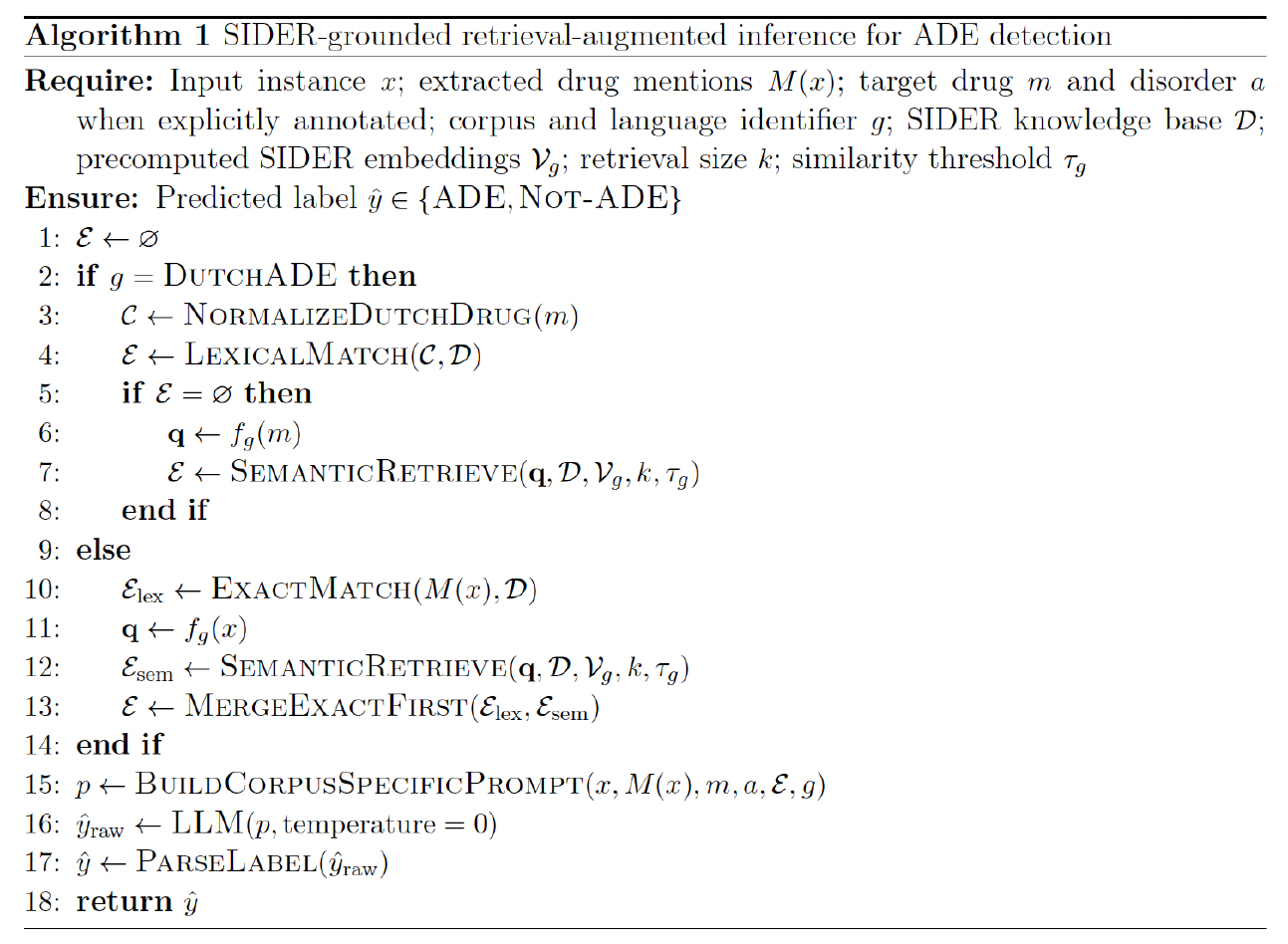
Extracted drug mentions are matched lexically and semantically against a SIDER-based knowledge base, with a sequential lexical-first branch for Dutch ADE notes and a parallel lexical+semantic branch for English and multilingual corpora. Retrieved evidence is inserted into a corpus-specific prompt, and a frozen Llama-3.1 model predicts a binary `ADE/Not-ADE` label under deterministic decoding.

Our results reveal corpus-dependent differences in ADE discourse, knowledge-base alignment, and detection performance. The topic analysis shows that patient-generated, clinical, and scientific sources differ in the adverse experiences they foreground and in the level of thematic specificity with which those experiences are described. The retrieval analysis reveals uneven alignment between real-world drug mentions and SIDER’s standardized vocabulary, with coverage ranging from 9.5% in patient-forum narratives to 91.0% in biomedical literature. Detection performance also varies by corpus, with no single configuration dominating across all five settings (Section 4.1). Together, these findings demonstrate that data-production context shapes what ADE evidence is expressed, how it is framed and standardized, and how readily it can be retrieved and computationally detected.

## 2. Background and Related Work

This section reviews two areas relevant to this study: variation in pharmacovigilance evidence across data-production contexts, and computational approaches—detection models and knowledge-grounded retrieval—that depend on corpus-specific supervision or structured pharmacovigilance knowledge.

### ADE Evidence Across Data-Production Contexts

Pharmacovigilance increasingly draws on evidence from patient-generated content, electronic health records (EHRs), medical case reports, biomedical literature, and curated drug-safety resources. These sources are complementary but not interchangeable. Patient forums and social media platforms provide direct accounts of medication use and adverse experiences, often expressed in colloquial, subjective, and multilingual language. Comparative reviews suggest that patient-generated sources can supplement spontaneous reports, product labels, and clinical literature by contributing timely or unexpected adverse-event evidence and information about how adverse experiences affect patients’ daily lives.^9^ At the same time, such data exhibit substantial linguistic variability, incomplete context, and platform-specific biases. Clinical notes contain longitudinal observations and documentation of suspected ADEs, but relevant evidence is often expressed indirectly in unstructured text and varies across institutions and documentation practices.^9^ Medical case reports and biomedical publications generally use more formal terminology and more explicit drug–outcome relations, but they are also shaped by clinical selection and scientific reporting practices.^21,25^ Consequently, each data-production context makes different aspects of drug-related harm visible and available for computational analysis.

Computational content analysis has also been used to characterize how patients discuss health experiences and medication use. Wexler *et al*. applied large-scale content analysis to online patient forums to identify recurring concerns and patterns in patient-generated health discourse,^11^ and Chen *et al*. combined adverse-effect extraction, signal detection, and topic modeling to examine patient narratives concerning methylphenidate, showing that topic analysis can surface contextual information about adverse effects, treatment practices, and misuse not captured by event extraction alone.^12^ Embedding-based approaches such as BERTopic have since enabled semantically related documents to be organized within a shared topic space.^26^ However, prior discourse analyses have generally focused on a single platform, patient population, or medication. Relatively little work has compared ADE discourse across patient-generated, clinical, case-report, and biomedical-literature sources or examined how these differences relate to alignment with curated pharmacovigilance knowledge and down-stream detection performance—the gap this study addresses.

### Computational Approaches to Detection and Knowledge Grounding

ADE detection has progressed from conventional classifiers such as support vector machines^28,29^ to transformer-based models, including BERT,^30^ Bio-ClinicalBERT,^31^ and RoBERTa,^32^ and more recently to LLMs adapted through prompting, fine-tuning, and data augmentation.^10,17,33^ Domain-adapted models often perform well when trained and evaluated within a single corpus,^34,35^ but Dai *et al*. showed through the MultiADE benchmark that models trained on one ADE domain did not generalize reliably to unseen text types,^23^ underscoring the influence of source language, annotation practices, and task formulation. Most LLM-based studies, however, remain limited to one or two source types and rarely examine source-specific supervision and structured external knowledge within the same cross-source analysis—a gap we address by evaluating prompting, QLoRA fine-tuning, and SIDER-grounded inference across five heterogeneous corpora.

Two adaptation strategies dominate this space and rely on different representations of pharmacovigilance evidence. Parameter-efficient fine-tuning methods such as Low-Rank Adaptation (LoRA) and Quantized LoRA (QLoRA) freeze most pretrained parameters and update only a small set of low-rank weights, learning from local, corpus-specific annotations while reducing memory requirements.^36^ Shakya *et al*. and Estupiñán-Ojeda *et al*. both found that QLoRA approaches full fine-tuning performance—on clinical summarization and bilingual primary-care notes, respectively—with substantially fewer trainable parameters and lower memory use,^27,36^ supporting QLoRA as a practical approach for learning source-specific linguistic patterns. Retrieval-augmented generation (RAG), by contrast, grounds inference in external evidence without updating model parameters, and depends instead on the coverage and terminology of the external resource. MALADE orchestrates LLM agents over drug labels, biomedical literature, and FDA resources to extract structured drug–outcome associations;^37^ Nygren *et al*. applied dense retrieval over SIDER for drug–side-effect queries;^15^ and AskFDALabel retrieves FDA labeling content for adverse-event annotation.^13^ These studies demonstrate the value of pharmacovigilance-grounded inference but focus primarily on querying or structured association extraction rather than ADE detection across heterogeneous text sources. We examine how these two forms of adaptation—local supervision versus external knowledge grounding—behave across multiple data-production contexts, and how variation in SIDER coverage specifically relates to detection performance.

## 3. Methods

We combine cross-source discourse analysis with ADE detection experiments to examine how data-production context shapes both the expression of adverse drug events and the performance of computational detection methods. Figure 1 summarizes the overall study design. Panel 1 introduces the five ADE corpora, which represent distinct languages and data-production settings, and shows their preprocessing into a common binary ADE detection task. Panel 2 presents a cross-source analysis of how adverse drug experiences are discussed in patient-generated text, medical case reports, clinical notes, and biomedical literature. Panel 3 evaluates corpus-specific QLoRA fine-tuning, zero- and few-shot prompting, and SIDER-grounded retrieval-augmented inference for ADE detection, together with instance-level SIDER retrieval coverage as a measure of how well real-world drug mentions align with standardized pharmacovigilance knowledge.

### 3.1. Data Sources and Task Construction

We use five expert-annotated ADE corpora spanning patient-generated, clinical, and scientific data-production contexts. Because the corpora differ in annotation schemes and units of analysis (sentence-, document-, and relation-level), we map their outputs to a shared binary label space (`ADE`, `Not-ADE`) while retaining corpus-specific operational criteria. Performance is therefore evaluated within each corpus.

**ADE Corpus V2** comprises medical case reports with sentence-level ADE annotations (17,637 train, 5,879 test). Following reports of cross-split duplication,^19^ we normalize case and whitespace and remove exact train–test duplicates.

**SMM4H-2026 Task 1** addresses document-level ADE detection in multilingual user-generated health content (German, French, Russian, English, Mandarin, Japanese), using predefined training, validation, and test splits with normalized formatting.

**CADEC V2** extends the AskAPatient corpus with drug and adverse-reaction annotations. We convert `brat` annotations to sentence-level binary labels and construct document-level splits to prevent sentences from the same review appearing across partitions.

**The Dutch ADE Corpus** consists of ICU clinical notes annotated with drug–disorder relations. We map adverse drug events to `ADE` and indications to `Not-ADE`, and split the 102 notes at the note level (70/15/15; seed 922) to prevent leakage.

**TwiMed** contains PubMed sentences and Twitter posts annotated for drugs and outcomes; we use the PubMed subset only. Sentences containing at least one outcome-negative relation are labeled `ADE`, and we construct a stratified 80/20 train–test split.

### 3.2. Cross-Genre ADE Discourse via Topic Modelling

To examine how ADE discourse varies across data-production contexts, we focus on antihypertensives and antidepressants, two widely used drug classes associated with diverse adverse effects and represented across multiple corpora. The analysis includes ADE-positive English-language texts from ADE Corpus V2, CADEC V2, TwiMed-PubMed, and the English subset of SMM4H-2026 Task 1.

Drug mentions are obtained from gold-standard entity annotations for CADEC V2 and TwiMed-PubMed. Because ADE Corpus V2 and SMM4H-2026 do not provide complete drug-entity annotations for the classification instances, mentions are extracted using Llama-3.1-405B and verified against the original text. The Dutch ADE Corpus is excluded from this analysis because its Dutch-language texts are not directly comparable within the shared English-language topic space and provide limited representation of the selected drug classes; it remains included in the SIDER-coverage and ADE-detection analyses.

ADE-positive instances mentioning either drug class are pooled across the four corpora. To reduce clustering driven by medication names rather than adverse experiences, extracted drug mentions are replaced with a generic placeholder prior to embedding. A single BERTopic model^26^ is then fitted to the combined drug-masked texts, placing all instances in a shared topic space. BERTopic generates sentence-level embeddings, applies UMAP for dimensionality reduction and HDBSCAN for clustering, and derives topic representations using class-based TF-IDF. For each topic, we inspect the highest-weighted terms and representative sentences and assign a descriptive label. Topic prevalence within each corpus is calculated as the proportion of qualifying instances assigned to that topic. We then compare prevalence, representative terms, and example sentences across sources to identify shared themes, source-concentrated concerns, and differences in how similar adverse experiences are framed. Because corpus sizes differ substantially, we interpret topic prevalence comparatively rather than as a direct estimate of population frequency.

### 3.3. Adverse Drug Event Detection Approaches

We evaluate three ADE detection settings: zero- and few-shot prompting, corpus-specific QLoRA fine-tuning, and SIDER-grounded retrieval-augmented inference. All configurations use corpus-specific instructions reflecting the unit of analysis and operational ADE criteria.

#### Zero- and Few-Shot Inference

Llama-3.2-3B-Instruct, Llama-3.1-70B-Instruct, and Llama-3.1-405B-Instruct are evaluated under zero-shot prompting. Each prompt includes the corpus-specific task definition, the input (sentence, document, or drug–disorder context), and an instruction to return one of two labels: `ADE` or `Not-ADE` . Llama-3.1-70B-Instruct and Llama-3.1-405B-Instruct are additionally evaluated under few-shot prompting with labeled demonstrations. These baselines do not include retrieved SIDER evidence.

#### QLoRA Fine-Tuning

We fine-tune a separate Llama-3.2-3B-Instruct model for each corpus using Quantized Low-Rank Adaptation (QLoRA).^38^ The base model is loaded with 4-bit NormalFloat quantization (double quantization), with computation in `bfloat16` precision. Following standard LoRA parameterization, low-rank adapters (rank *r* = 16, scaling *α* = 32, dropout 0.05) are applied to all linear layers; base-model parameters remain frozen and only adapter weights are updated. Each training instance is formatted as a corpus-specific instruction-following exchange using the Llama chat template, where the system message defines the task, the user message contains the input, and the assistant message provides the gold label (`ADE` or `Not-ADE`). Models are trained with a causal language-modeling objective using a paged 8-bit AdamW optimizer, cosine learning-rate schedule, and gradient checkpointing.^38^ At inference, the assistant response is omitted, greedy decoding is applied, and outputs are mapped to the binary label space.

#### SIDER-Grounded Retrieval-Augmented Inference

Our retrieval-augmented configuration introduces external pharmacovigilance evidence without task-specific parameter updates. Llama-3.1-70B-Instruct and Llama-3.1-405B-Instruct are served with vLLM and remain frozen during inference. Each input is augmented with evidence from SIDER 4.1, a curated resource containing drug–adverse-effect associations derived primarily from drug labels and clinical-trial information, with adverse effects standardized using MedDRA terminology.^14^

We constructed a knowledgebase mapping canonical drug names from SIDEr and synonyms to lists of associated adverse effects (re-stricted to MedDRA Pre-ferred Terms), retaining up to ten effects per entry to limit prompt length. Very short drug names are excluded from the lexical index to reduce ambiguity.

Retrieval combines exact lexical matching with embedding-based semantic retrieval and is adapted to the language and annotation structure of each corpus. For English-language inputs, lexical matching is applied to extracted drug mentions *M* (*x*), while semantic re-trieval uses the full input text as a query. The two evidence sources are merged, prioritizing exact matches when duplicates occur. Semantic retrieval uses `BAAI/bge-large-en-v1.5` for English inputs and `paraphrase-multilingual-MiniLM-L12-v2` for cross-lingual inputs, retaining the top *k* = 3 entries above corpus-specific similarity thresholds (*τ*_en_ = 0.80, *τ*_nl_ = 0.62).

For the Dutch ADE Corpus, retrieval follows a sequential strategy: the target drug entity is first normalized using rule-based transformations to approximate English forms (e.g., `-ine` →`-in`, `-ole` →`-ol`), and lexical matching is attempted before falling back to cross-lingual semantic retrieval if needed. The SMM4H corpus uses the English retrieval branch for English-language instances and the cross-lingual branch otherwise.

Retrieved evidence is inserted into a corpus-specific prompt containing the original input and any available drug or disorder. The model is instructed to use the retrieved information as supporting evidence and to generate `ADE` or `Not-ADE` . Inference uses deterministic decoding (temperature 0), and outputs are normalized and mapped to the binary label space. We evaluate both exact-match-only and hybrid lexical–semantic retrieval settings where applicable.

## 4. Results

### SIDER Coverage Across Data-Production Contexts

Table 1 reports instance-level SIDER retrieval coverage across the five corpora. Exact coverage indicates that at least one identified drug mention matched a canonical drug name or synonym in SIDER. Semantic-only coverage indicates that a SIDER entry was retrieved through embedding similarity when no exact match was available. Total coverage represents retrieval through either route.

**Table 1:** Instance-level SIDER retrieval coverage across corpora. Semantic-only coverage denotes evidence retrieved through embedding similarity in the absence of an exact lexical match.

| Corpus | Exact (%) | Semantic-only (%) | Total (%) |
| --- | --- | --- | --- |
| ADE Corpus V2 | 52.0 | 0.0 | 52.0 |
| SMM4H-2026 | 4.7 | 58.3 | 63.0 |
| Dutch ADE | 28.2 | 2.5 | 30.6 |
| CADEC V2 | 9.5 | 0.0 | 9.5 |
| Twimed-PubMed | 91.0 | 0.0 | 91.0 |

SIDER coverage varied substantially across data-production contexts. TwiMed-PubMed achieved the highest total coverage at 91.0%, almost entirely through exact matching, consistent with the formal and standardized medication terminology used in biomedical literature. ADE Corpus V2 showed moderate exact-match coverage of 52.0%, while the Dutch ADE Corpus achieved 30.6% total coverage, including a modest 2.5 percentage-point contribution from cross-lingual semantic retrieval.

The largest semantic contribution occurred in SMM4H-2026. Exact matching covered only 4.7% of instances, whereas semantic retrieval increased total coverage to 63.0%. This pattern reflects the multilingual and linguistically variable nature of user-generated health content, for which direct correspondence with SIDER’s predominantly English and standardized vocabulary is limited. Because semantic retrieval uses the full input text, this coverage represents retrieval of related SIDER evidence rather than necessarily establishing a direct lexical link between the reported drug mention and a SIDER entry.

CADEC V2 showed the lowest total coverage, with 9.5% of instances receiving exact SIDER evidence and no additional entries passing the semantic threshold. Examination of unmatched mentions showed recurrent mismatches involving brand names, spelling variation, abbreviations, and colloquial forms. These findings demonstrate that the availability of curated pharmacovigilance evidence depends strongly on the terminology and documentation practices of each source; in particular, high coverage in formal biomedical text does not necessarily translate to patient-generated or multilingual settings. In the detection analysis below, these coverage differences help explain where retrieval-based configurations are most competitive.

### Cross-Genre Patterns in ADE Discourse

Figures 3 and 4 summarize the pooled BERTopic analysis of 6,713 drug-masked, ADE-positive instances from the four English-language corpora, comprising 6,001 instances from CADEC V2, 466 from SMM4H-2026, 181 from ADE Corpus V2, and 65 from TwiMed-PubMed. The model identifies nine interpretable topics in addition to an outlier cluster. Sexual side effects, neurological symptoms, and swelling or localized pain appear across multiple corpora, whereas suicidal or panic-related thoughts, night sweats, vivid dreams, and memory difficulties are concentrated in CADEC V2. These patterns suggest that long-form patient narratives provide greater visibility into emotionally salient and highly specific experiences that are rarely represented in shorter or more formally structured sources.

**Fig. 3:**
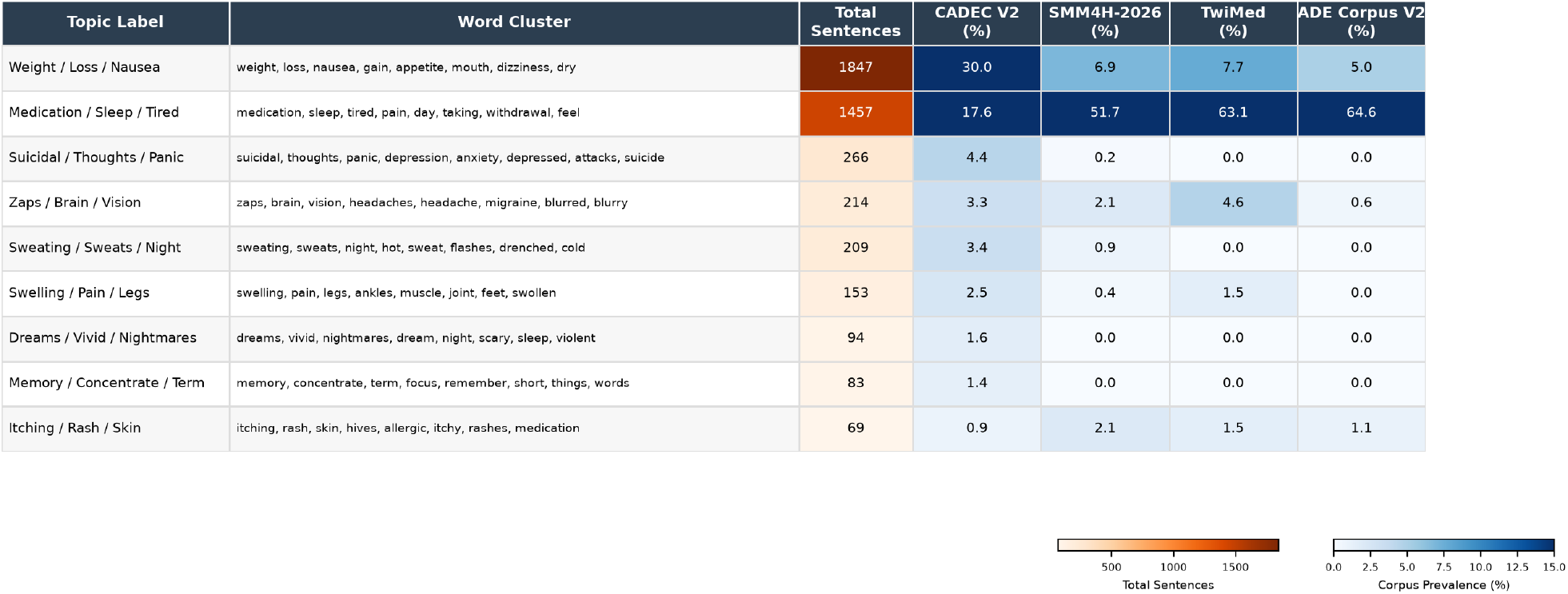
Cross-genre ADE discourse topics identified by BERTopic (antihypertensive + antidepressant case study, CADEC V2, SMM4H-2026, TwiMed, ADE Corpus V2).

**Fig. 4:**
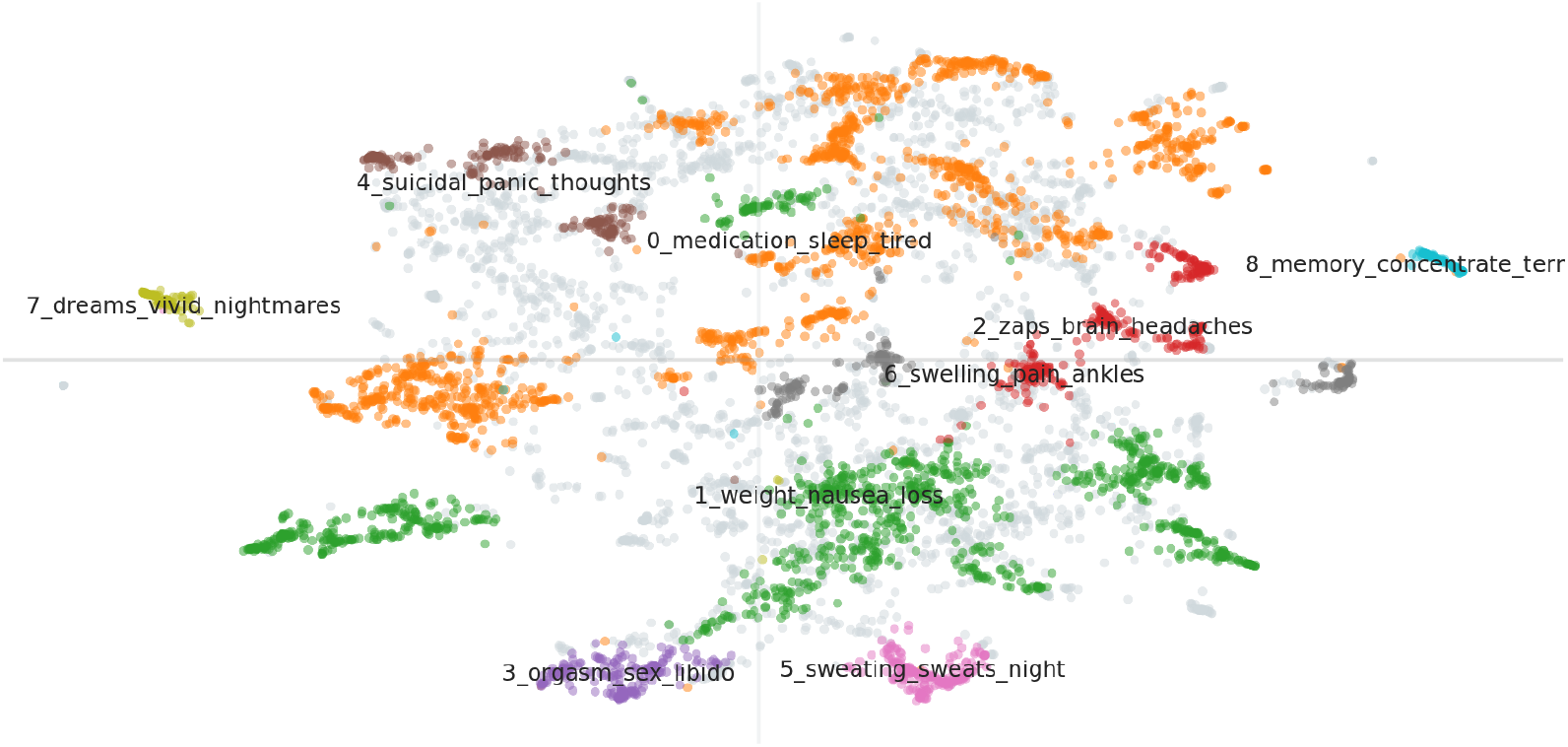
UMAP projection of drug-masked ADE-positive instances colored by BERTopic topic assignment. Gray points denote outliers. Topics with distinctive vocabularies form tight clusters, whereas the two largest, more generic topics (Medication/Sleep/Tired/Pain and Weight/Nausea/Loss/Gain) occupy a more diffuse region of the embedding space.

Although *Weight/Loss/Nausea* is the largest topic in the pooled corpus, *Medication/Sleep/Tired* dominates SMM4H-2026, TwiMed-PubMed, and ADE Corpus V2, accounting for 51.7%, 63.1%, and 64.6% of their qualifying instances, respectively. In contrast, CADEC V2 is distributed across a broader range of symptom-specific topics, with *Weight/Loss/Nausea* accounting for 30.0% and *Medication/Sleep/Tired* for 17.6%. Because CADEC V2 contributes most of the pooled instances, prevalence estimates for ADE Corpus V2 and TwiMed-PubMed should be interpreted descriptively rather than as precise frequency estimates. Consistent with this pattern, Figure 4 shows that topics with more distinctive vocabularies form relatively compact clusters, whereas the two largest and more general topics occupy more diffuse regions of the embedding space.

Topic prevalence is broadly similar across antidepressant- and antihypertensive-related instances, supporting their representation within a shared topic space. The most notable differences are clinically plausible: swelling and ankle pain are more prevalent among antihypertensive-related instances, whereas sexual side effects are more prevalent among antidepressant-related instances. Overall, the findings indicate that patient forums provide non-redundant pharmacovigilance evidence, particularly for subjective, psychologically salient, and highly specific adverse experiences that are less visible in social-media posts, medical case reports, and biomedical literature. From a sociotechnical perspective, these differences reflect how platform structure, documentation purpose, intended audience, and opportunities for extended narration shape which adverse experiences become visible for computational analysis.

### Detection Performance Across Corpora

Table 2 compares zero-shot, few-shot, QLoRA, and RAG-SIDER configurations across the five corpora. Because the datasets are generally imbalanced toward `Not-ADE`, we report *F*1_micro_ together with ADE-class precision and recall to distinguish overall classification performance from performance on the clinically relevant ADE class. QLoRA-Llama-3.2-3B achieves the highest *F*1_micro_ on ADE Corpus V2, SMM4H-2026, and CADEC V2 (0.91, 0.80, and 0.88, respectively). These three corpora provide more labeled training examples than the Dutch ADE Corpus and TwiMed-PubMed, suggesting that corpus-specific fine-tuning is particularly effective when sufficient in-domain supervision is available. In these settings, the compact QLoRA-adapted model also matches or exceeds the performance of substantially larger inference-only models.

**Table 2:**
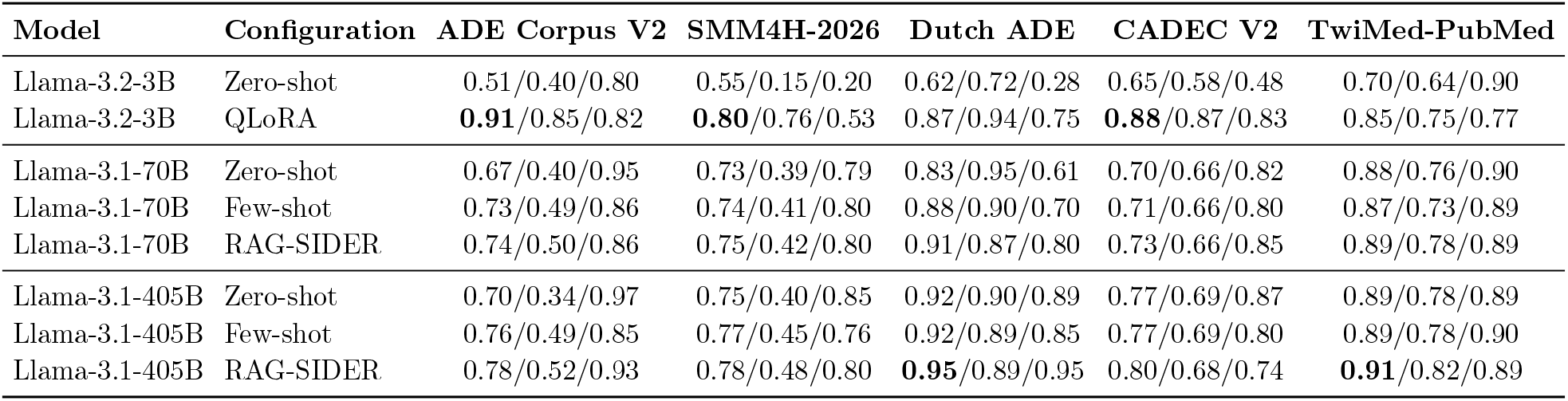
ADE detection performance across five corpora. Each cell reports *F* 1_micro_*/P*_ADE_*/R*_ADE_. Within each corpus, the highest *F* 1_micro_ is shown in bold; values are intended as descriptive rather than a controlled comparison across model families.

RAG-SIDER with Llama-3.1-405B attains the highest *F* 1_micro_ on the Dutch ADE Corpus and TwiMed-PubMed (0.95 and 0.91), and on the Dutch ADE Corpus achieves an ADE-class recall of 0.95, indicating high sensitivity to positive ADE instances. Few-shot prompting yields inconsistent changes relative to zero-shot prompting across corpora, with clear improvements in some settings and minimal or no benefit in others.

Overall, the strongest configuration is corpus-dependent: QLoRA performs best on the three corpora with greater task-specific supervision, whereas large-model RAG-SIDER is most competitive on the two smaller corpora where alignment with SIDER is higher. These patterns should not be interpreted as a controlled comparison of adaptation strategies, because the configurations differ in model scale, supervision, and retrieval evidence. Direct comparison with prior work is also limited by differences in task formulation and splits (for example, our ADE Corpus V2 experiments use a leakage-corrected split, and the Dutch ADE Corpus and CADEC V2 lack established binary baselines). We therefore emphasize comparisons among configurations evaluated on the same held-out test set within each corpus.

## 5. Discussion and Limitations

This study shows that data-production context shapes which adverse drug experiences are expressed, how they are framed, and how readily they align with curated pharmacovigilance resources. Patient forums, social-media posts, case reports, EHR notes, and biomedical literature are therefore not interchangeable sources of ADE evidence.

SIDER coverage ranges from 91.0% exact matches in TwiMed-PubMed to 9.5% in CADEC V2, reflecting the extent to which SIDER encodes institutional conventions—English, generic drug names, standardized terminology—that mirror biomedical literature more than patient-generated or multilingual content. These gaps imply that systems relying heavily on SIDER-like resources may overrepresent clinically formal sources and underrepresent patients’ language, making coverage a sociotechnically meaningful property, not just a technical metric.

Topic analysis indicates that CADEC V2 foregrounds subjective, symptom-specific, and psychologically salient themes (e.g., sexual side effects, suicidal or panic-related thoughts, vivid dreams), whereas social media, case reports, and biomedical literature concentrate on broader medication-, sleep-, tiredness-, and pain-related discourse. Patient forums thus provide non-redundant pharmacovigilance evidence shaped by platform structure, audience, and documentation purpose.

Detection results are descriptive probes of these representational differences. QLoRA-Llama-3.2-3B performs best in corpora with greater in-domain supervision, whereas RAG-SIDER with Llama-3.1-405B is most competitive where SIDER coverage is higher (Dutch ADE, TwiMed-PubMed). No single adaptation strategy dominates across contexts, suggesting that model design should be conditioned on both supervision and knowledge-base alignment rather than optimized globally.

Our comparison confounds model scale and training-set size, relies on binary ADE detection with single-run evaluations, and uses topic models dominated by CADEC V2 and LLM-based drug-mention extraction in some corpora; topic and coverage estimates should therefore be interpreted descriptively. Future work should use matched model scales, repeated runs, joint extraction and normalization pipelines, and multimodal or workflow-linked datasets. Practically, combining source-sensitive detectors with knowledge-grounded retrieval, while routinely auditing coverage and alignment, may help pharmacovigilance pipelines remain responsive both to formally documented harms and to adverse experiences articulated by patients in their own words.

## Data Availability

All data produced are available online.
https://data.csiro.au/collection/csiro:62387
https://huggingface.co/datasets/SetFit/ade_corpus_v2_classification

https://uvaauas.figshare.com/articles/dataset/Dutch_ADE_corpus/28846991

https://huggingface.co/datasets/SetFit/ade_corpus_v2_classification

https://data.csiro.au/collection/csiro:62387

## Notes

### Competing Interest Statement

The authors have declared no competing interest.

### Author Declarations

Our study analyzed record-level textual data rather than aggregated or summary data. We used five previously released research corpora: ADE Corpus V2: sentences from published MEDLINE case reports; Dutch ADE Corpus: anonymized clinical notes; CADEC v2: patient-authored posts from a public medical forum; TwiMed: text from PubMed and public social-media posts; and SMM4H: public social-media posts. The Dutch clinical notes were anonymized by the dataset creators before we received them. Any direct identifiers in the social-media data were removed or replaced before our analysis.

## References

1. M. S. Donaldson, J. M. Corrigan and L. T. Kohn, To err is human: building a safer health system (2000).

2. X. Qiu, S. Shao, H. Wang and X. Tan, Bio-k-transformer: A pre-trained transformer-based sequence-to-sequence model for adverse drug reactions prediction, Computer Methods and Programs in Biomedicine 260, p. 108524 (2025).

3. D. C. Classen, S. L. Pestotnik, R. S. Evans, J. F. Lloyd and J. P. Burke, Adverse drug events in hospitalized patients: excess length of stay, extra costs, and attributable mortality, Jama 277, 301 (1997).

4. M. De Rosa, G. Fenza, A. Gallo, M. Gallo and V. Loia, Pharmacovigilance in the era of social media: discovering adverse drug events cross-relating twitter and pubmed, Future Generation Computer Systems 114, 394 (2021).

5. E. Kopacheva, A. Henriksson, H. Dalianis, T. Hammar and A. Lincke, Identifying adverse drug events in clinical text using fine-tuned clinical language models: Machine learning study, JMIR Formative Research 9, p. e71949 (2025).

6. S. Golder, K. O’Connor, G. Lopez-Garcia, N. P. Tatonetti and G. Gonzalez-Hernandez, Leveraging unstructured data in electronic health records to detect adverse events from pediatric drug use: A scoping review, Annual review of biomedical data science 8, 227 (2025).

7. L. Chen, Y. Gu, X. Ji, Z. Sun, H. Li, Y. Gao and Y. Huang, Extracting medications and associated adverse drug events using a natural language processing system combining knowledge base and deep learning, Journal of the American Medical Informatics Association 27, 56 (2020).

8. I. Guellil, Y. Berrachedi, N. E. Chenni, M.-N. Abboud, J. Wu, H. Wu and B. Alex, Detecting adverse drug events in social media: A brief literature review, SN Computer Science 7, p. 199 (2026).

9. S. Golder, K. O’Connor, Y. Wang, A. Klein and G. Gonzalez Hernandez, The value of social media analysis for adverse events detection and pharmacovigilance: scoping review, JMIR public health and surveillance 10, p. e59167 (2024).

10. O. Elbiach, H. Grissette et al., Benchmarking large language models for adverse drug reaction extraction in social media and clinical texts, Results in Engineering, p. 107362 (2025).

11. A. Wexler, A. Davoudi, D. Weissenbacher, R. Choi, K. O’Connor, H. Cummings and G. Gonzalez-Hernandez, Pregnancy and health in the age of the internet: A content analysis of online “birth club” forums, PloS one 15, p. e0230947 (2020).

12. X. Chen, C. Faviez, S. Schuck, A. Lillo-Le-Louët, N. Texier, B. Dahamna, C. Huot, P. Foulquié, S. Pereira, V. Leroux et al., Mining patients’ narratives in social media for pharmacovigilance: adverse effects and misuse of methylphenidate, Frontiers in pharmacology 9, p. 541 (2018).

13. L. Wu, H. Fang, Y. Qu, J. Xu and W. Tong, Leveraging fda labeling documents and large language model to enhance annotation, profiling, and classification of drug adverse events with askfdalabel: L. wu et al., Drug Safety 48, 655 (2025).

14. M. Kuhn, I. Letunic, L. J. Jensen and P. Bork, The sider database of drugs and side effects, Nucleic acids research 44, D1075 (2016).

15. S. Nygren, O. Erdogan, P. Avci, A. Daniels, R. Rassool, A. Beheshti and D. Galeano, Rag-based architectures for drug side effect retrieval using compact llms, Scientific Reports 16, p. 12754 (2026).

16. H. Prioleau, S. K. Aryal and J. Blackstone, Leveraging large language models for adverse drug event detection: A comparative study of token and span-based named entity recognition, 205 (2025).

17. F. Zeidi, R. Christof, R. König and L. Childs, ‘pei at# smm4h-heard 2025: Enhancing adverse event detection via error-driven data augmentation, p. 56 (2025).

18. Z. Duan, K. Wei, Z. Xue, J. Zhou, S. Yang, S. Ma, J. Jin et al., Crowdsourcing-based knowledge graph construction for drug side effects using large language models with an application on semaglutide, arXiv preprint arXiv:2504.04346 (2025).

19. Z. Huang, J. Lu, Q. Sun and H. Lu, Multi-axis evaluation of adverse drug event classification: Clinical error cost, linguistic failure taxonomy, and leakage-corrected benchmarks on ade corpus v2, 223 (2026).

20. F. Dong, W. Guo, J. Liu, T. A. Patterson and H. Hong, Bert-based language model for accurate drug adverse event extraction from social media: implementation, evaluation, and contributions to pharmacovigilance practices, Frontiers in Public Health 12, p. 1392180 (2024).

21. H. Gurulingappa, A. M. Rajput, A. Roberts, J. Fluck, M. Hofmann-Apitius and L. Toldo, Development of a benchmark corpus to support the automatic extraction of drug-related adverse effects from medical case reports, Journal of biomedical informatics 45, 885 (2012).

22. G. Lopez-Garcia, J. Cortina, J. Berkowitz, J. Chan, S. K. Dey, I. F. Amaro, F. Gallego, L. Gryboski, A. Z. Klein, F. Z. Kolehparcheh et al., Overview of the 11th social media mining for health (# smm4h) and health real-world data (heard) shared tasks at acl 2026, 353 (2026).

23. X. Dai, S. Karimi, A. Sarker, B. Hachey and C. Paris, Multiade: A multi-domain benchmark for adverse drug event extraction, Journal of Biomedical Informatics 160, p. 104744 (2024).

24. R. M. Murphy, D. A. Dongelmans, N. F. de Keizer, R. J. Jongeneel, C. H. Koster, K. J. Jager Abu-Hanna, I. Calixto and J. E. Klopotowska, Creation of a gold standard dutch corpus of clinical notes for adverse drug event detection: the dutch ade corpus, Language Resources and Evaluation 59, 2763 (2025).

25. N. Alvaro, Y. Miyao and N. Collier, Twimed: Twitter and pubmed comparable corpus of drugs, diseases, symptoms, and their relations, JMIR public health and surveillance 3, p. e24 (2017).

26. M. Grootendorst, Bertopic: Neural topic modeling with a class-based tf-idf procedure, arXiv preprint arXiv:2203.05794 (2022).

27. P. R. Shakya, A. Khaneja and K. B. Wagholikar, For clinical data extraction, qlora attains accuracy close to lora while requiring lower compute resources, medRxiv, 2025 (2025).

28. J. Liu, G. Wang and G. Chen, Identifying adverse drug events from social media using an improved semisupervised method, IEEE Intelligent Systems 34, 66 (2019).

29. J. Wang, L. Zhao, Y. Ye and Y. Zhang, Adverse event detection by integrating twitter data and vaers, Journal of biomedical semantics 9, p. 19 (2018).

30. J. Devlin, M.-W. Chang, K. Lee and K. Toutanova, Bert: Pre-training of deep bidirectional transformers for language understanding, 1, 4171 (2019).

31. E. Alsentzer, J. Murphy, W. Boag, W.-H. Weng, D. Jindi, T. Naumann and M. McDermott, Publicly available clinical bert embeddings, 72 (2019).

32. Y. Liu, M. Ott, N. Goyal, J. Du, M. Joshi, D. Chen, O. Levy, M. Lewis, L. Zettlemoyer and V. Stoyanov, Roberta: A robustly optimized bert pretraining approach, arXiv preprint arXiv:1907.11692 (2019).

33. H. Li and H. Lin, Adverse drug reaction detection on social media based on large language models, Information 17, p. 352 (2026).

34. C. Hiba, E. H. Nfaoui and C. Loqman, Fine-tuning transformer models for adverse drug event identification and extraction in biomedical corpora: A comparative study, 957 (2023).

35. K. Huang, J. Altosaar and R. Ranganath, Clinicalbert: Modeling clinical notes and predicting hospital readmission, arXiv preprint arXiv:1904.05342 (2019).

36. C. Estupiñán-Ojeda, R. J. Sandomingo-Freire, L. Padró and J. Turmo, High-fidelity parameter-efficient fine-tuning for joint recognition and linking of diagnoses to icd-10 in non-standard primary care notes, JAMIA open 8, p. ooaf120 (2025).

37. J. Choi, N. Palumbo, P. Chalasani, M. M. Engelhard, S. Jha, A. Kumar and D. Page, Malade: Orchestration of llm-powered agents with retrieval augmented generation for pharmacovigilance, arXiv preprint arXiv:2408.01869 (2024).

38. P. N. Srinivasu, A. Samudrala, P. G. Devesh, S. A. Reddy and A. A. Nuaim, Quantized low-rank adaptation in large language models for clinical text simplification, International Journal of Computational Intelligence Systems 19, p. 267 (2026).

